# Association between DNMT3A-driven clonal hematopoiesis, trained immunity and immune cell function in obesity

**DOI:** 10.64898/2026.08.20.26360882

**Authors:** Harsh Bahrar, Helin Tercan, Benjamin C. Cossins, Nils Rother, Rosanne C van Deuren, Alexander Hoischen, Leo AB Joosten, Mihai G Netea, Siroon Bekkering, Niels P. Riksen

## Abstract

Trained immunity and clonal hematopoiesis are two newly identified immunological phenomena that contribute to the pathophysiology of atherosclerotic cardiovascular disease. These two phenomena share some convergent molecular mechanisms, such as IL-1β being a central regulator and involvement of epigenetic enzymes. Therefore, we hypothesize that presence of clonal hematopoiesis driver mutations (CHDMs) can predispose to an increased capacity to build trained immunity. We previously characterized how the presence of CHDMs relates to immune cell function and vasculometabolic complications in a cohort of older individuals with overweight and obesity. From this cohort we now selected 17 individuals with CH due to DNMT3A mutations and 15 without any known CHDMs. We performed in depth immune characterization via flow cytometry, functional assays with monocytes and neutrophils, and we measured the capacity to build trained immunity using β-glucan and oxLDL as stimuli. We corroborated our previous findings of lower ex vivo cytokine production capacity of PBMCs from individuals with DNMT3A mutations. Importantly, presence of DNMT3A CHDMs associated with higher trained immunity response. Moreover, we demonstrated that individuals with DNMT3A mutations were characterized with higher CD10^+^ mature neutrophils and a lower neutrophil MPO release upon TLR2 stimulation. In conclusion, presence of DNMT3A CHDMs is associated with increased susceptibility to build a hyperresponsive trained monocyte phenotype. The exact molecular mechanisms behind this phenomena requires further investigation.

## Introduction

Ageing gradually alters the composition and the function of the immune system. Adaptive immune response becomes impaired, marked by restricted T cell receptor repertoire and reduced antibody diversity^1^. Whereas innate immune cells display a heightened proinflammatory response on the baseline. Age-associated inflammation, also known as inflammaging, can lead to plethora of pathologies^2^. Clonal hematopoiesis is one of such age-associated phenomena^3^.

Clonal hematopoiesis is defined as clonal expansion of leukocytes due to somatic mutations in hematopoietic stem cells. These somatic mutations can confer a certain survival or fitness advantage, leading to clonal expansion of leukocytes with this mutation. The presence of a clonal hematopoiesis driver mutation (CHDM) in circulating leukocytes with a variant allele frequency (VAF) ≥2%, and without evidence of hematological malignancy, dysplasia, or cytopenia is defined as Clonal Hematopoiesis of Indeterminate Potential (CHIP)^4^. Many epidemiolocal studies have established that CHIP is associated with an increased risk of atherosclerotic cardiovascular disease (ASCVD)^5^, although we have previously reported that it is not associated with the presence of asymptomatic atherosclerotic plaques *per se*^6^. Recent work with increased sequencing depth showed that also CHDMs with VAF<2% can have clinical significance^7^. Mutations in the *DNMT3A* gene are the leading CHDMs in most cohorts, followed by TET2 and ASXL1 mutations^8^.

Clonal hematopoiesis is a risk factor for obesity-associated metabolic complications and for ASCVD^9^. Despite strong evidence for a correlation between clonal hematopoiesis and ASCVD, the causal mechanisms are not fully understood. TET2 deficient mouse models have been shown to display accelerated atherosclerosis, marked by increased NLRP3-inflammasome driven IL-1β production by macrophages^10^. A more recent study showed a convergent phenotype of atherosclerosis in DNMT3A deficient mice, with increased intracellular pro-IL-1β expression measured with flow cytometry^11^. Single cell RNA sequencing of isolated monocytes of six patients with heart failure and CHIP due to DNMT3A CHDMs revealed a proinflammatory RNA expression profile at baseline^12^. In contrast, we recently observed a lower cytokine production capacity of isolated PBMCs from individuals with obesity and CHIP, after *ex vivo* exposure to LPS, compared to individuals with obesity and without CHDMs^6^.

In the current study, we aim to explore how CHIP due to mutations in DNMT3A affects the phenotype of innate immune cells in more detail. In particular, we focus on trained immunity and on neutrophil function, as two novel protagonists of atherosclerosis pathophysiology. Trained immunity describes the persistent functional hyperresponsive phenotype of innate immune cells after brief stimulation to micro-organisms or to atherogenic stimuli, such as oxLDL^13^. A key driver of trained immunity, at least in the context of oxLDL^14^, and of β-glucan^15^ is IL-1β. Recent murine studies provided strong proof that trained immunity can accelerate atherosclerosis development, in the setting of hyperglycemia^16^, intermittent high-fat diet^17^, and post myocardial infarction^18^. Given the knowledge that CHDMs are mainly mutations in epigenetic enzymes and epigenetic reprogramming is the central mechanism of trained immunity, and cells with CHDMs produce more IL-1β, which can induce trained immunity, we hypothesize that the presence of CHDMs is associated with an increased potential to develop a trained immunity phenotype.

A second aim of this study is to explore the effects of CHDMs on neutrophil function. Most studies on CHIP focused on monocytes with only a few studies suggesting that CHDMs in TET2^19^ and JAK2^20^ can also affect neutrophil function. We recently showed that individuals with obesity and CHDMs have higher absolute neutrophil counts^6^. Accumulating evidence highlights the role of neutrophils in ASCVD^21,22^. Despite their short lifespan, neutrophils can also retain memory phenotype in their bone marrow progenitor cells^23^.

We previously identified CHDMs in a cohort of older individuals with overweight and obesity^6^. For the current study we included 17 individuals with DNMT3A mutations and 15 without any known CHDMs, matched on age, sex and BMI. We studied the immune phenotype of PBMCs, purified monocytes and neutrophils from these individuals in detail by characterizing surface marker expression by flow cytometry, leukocyte differentiation, and cytokine production capacity upon *ex vivo* stimulation. And lastly, we performed trained immunity experiments on purified monocytes.

## Methods

### Study design and population

Participants were selected from the “300-Obese (OB)” cohort based on presence or absence of DNMT3A mutations in the blood that was collected between 2014 and 2016, for simplicity we refer to this time point as 2016 sequencing. We also included the six individuals with a CHDM in TET2. We repeated DNA sequencing in 2023. Since the TET2 CHDM could not be replicated in three individuals and because our smMIP assay only covers a small part of TET2^6,7^, we decided to only focus on DNMT3A CHDMs in the current paper. For an overview of the CHDMs in 2016 and 2023, and how we defined the final groups, please refer to **Table 1** and Consort Diagram in **Figure 1**. For detailed description of the 300 OB cohort and the comprehensive characterization of CHDMs please refer to Tercan *et al*^6^. Comparable distribution of BMI, age and sex was considered when selecting the individuals. 32 individuals with a median BMI of 30 kg/m^2^, between the ages of 66 and 84, consisting of Western European ancestry, were recruited in the Radboud university medical center in 2021. The research protocol was approved by the Radboud University Ethical Committee (NL72552.091.20; 2020-6135), and all subjects gave written informed consent. The study protocol was performed in accordance with the 1975 Declaration of Helsinki.

**Figure 1:**
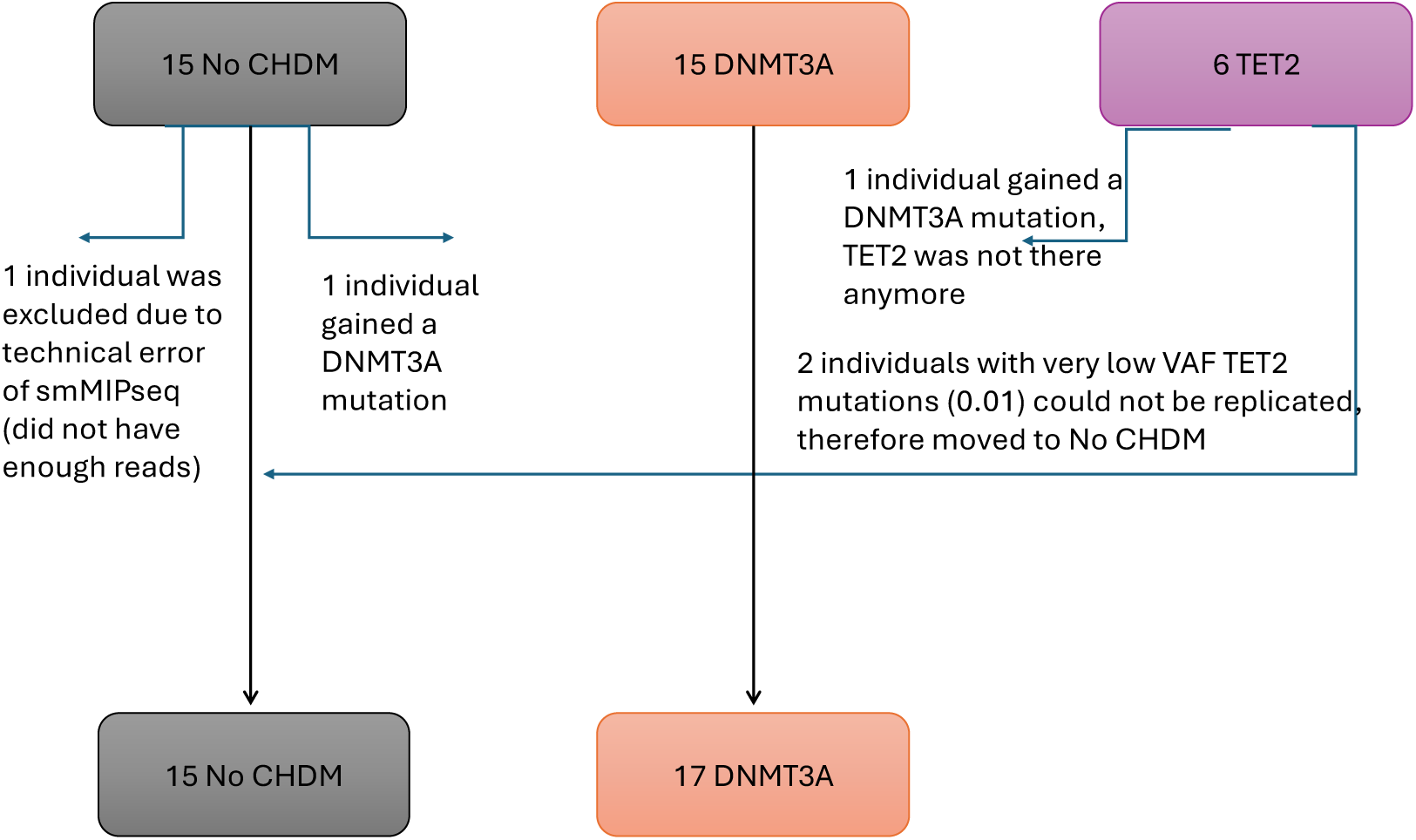
Consort diagram depicting the inclusion of individuals based on CHDMs identified in 2016 and 2023, final groups(No CHDM, DNMT3A) are included in the current study.

**Table 1:** Baseline characteristics.

|  | No CHDM (n=15) | DNMT3A (n=17) | Total (n=32) |
| --- | --- | --- | --- |
| Age (year) | 76 (75-77) | 73 (69-78) | 75 (72-77) |
| Sex (male, %) | 47 | 47 | 47 |
| Ancestry (Western European, %) | 100 | 100 | 100 |
| BMI (kg/m<sup>2</sup>) | 30 (29-31) | 30 (28-32) | 30 (29-32) |
| Waist circumference (cm) | 108 (103-111) | 115 (108-117) | 110 (106-117) |
| Systolic blood pressure (mmHg) | 148 (139-157) | 144 (142-152) | 144 (140-154) |
| Diastolic blood pressure (mmHg) | 84 (72-88) | 76 (72-82) | 78 (72-86) |
| Heart rate (beats per minute) | 64 (60-74) | 72 (60-72) | 72 (60-73) |
| Packyears (year) | 14 (10-28) | 17 (7-20) | 145 (8-24) |
| Diabetes Mellitus (Type II, %) | 7 | 6 | 6 |
| Lipid lowering drug use (%) | 70 | 80 | 75 |
| Total cholesterol (mmol/l) | 5.2 (4.8-5.9) | 4.9 (4.6-6) | 5.1 (4.7-6) |
| LDL cholesterol (mmol/l) | 3.1 (2.7-3.6) | 3.1 (2.5-3.8) | 3.1 (2.6-3.8) |
| Triglycerides (mmol/l) | 1.3 (1.1-1.6) | 1.5 (1.1-1.7) | 1.4 (1.1-1.6) |
| HDL cholesterol (mmol/l) | 1.4 (1.3-1.7) | 1.3 (1.2-1.5) | 1.3 (1.2-1.6) |
| Non-HDL cholesterol (mmol/l) | 3.8 (3.2-4.4) | 3.7 (3.2-4.3) | 3.8 (3.2-4.4) |
| Creatinine (umol/l) | 81 (69-89) | 79 (68-93) | 80 (68-90) |
| eGFR (ml/min/1.73 m<sup>2</sup>) | 72 (67-78) | 73 (69-80) | 73 (68-78) |
| Glucose (mmol/l) | 5.4 (5.2-6.2) | 5.6 (4.9-6.2) | 5.5 (5.2-6.2) |
| Hba1c (mmol/ mol) | 40 (40-42) | 38 (37-42) | 40 (38-42) |

Data shown as median (interquartile range 1-3) and percentage (%) where appropriate. BMI indicates body mass index; HDL, high-density lipoprotein; Hba1c Hemoglobin A1c

For each participant venous blood was collected between 8-9 am into BD Vacutainer® K2EDTA (10 ml) tubes. All laboratory procedures were performed immediately following the blood collection. When possible, patients with and without DNMT3A mutations were included in the same day.

### Blood sampling and chemistry parameters

2 EDTA vacutainer tubes were centrifuged for 10 minutes at 2749 g at RT to collect plasma. Plasma samples were then stored at –80°C until measurement.

Whole blood composition was assessed by a Sysmex-XN 450 hematology analyzer. Plasma hs-CRP, creatinine, total cholesterol, high-density lipoprotein (HDL) cholesterol, and triglyceride levels were determined by standard laboratory procedures. Low-density lipoprotein (LDL) cholesterol levels were calculated with the Friedewald equation.

### Identification of CHDMs

The first DNA samples were collected between 2014 and 2016 to identify presence of CHDMs. As mentioned above, individuals were selected for the current study based on these results. In the current study, we repeated DNA sequencing in 2023. CHDMs were identified in whole blood by the same ultra-sensitive assay as in the initial cross-sectional 300 OB cohort, as previously described^6,24^.

Briefly, 300 single-molecule molecular inversion probes (smMIP) were designed to cover a selection of well-known hotspots of a panel of 24 clonal hematopoiesis driver genes, including the entire DNMT3A gene. As the DNMT3A gene contains the most driver mutations and is shown to be causally linked to atherosclerosis in mouse models^11^, we only focused on CHDMs in this particular gene.

For each sample two technical polymerase chain reaction (PCR) replicates were run, thereafter two independent data processing strategies and a quality control step were performed. Variant allele frequencies were calculated using samtools mpileup^25^.

### Isolation of cells from peripheral blood

#### Peripheral Blood Mononuclear Cell (PBMC) isolation

PBMC isolation was performed with differential density centrifugation over Ficoll-Paque (GE Healthcare).

Briefly, blood was diluted in Phosphate Buffered Saline (PBS) (Gibco) and layered on Ficoll-Paque PLUS (Cytiva) density gradient centrifugation for 30 minutes at 615g (no brakes, RT).

The PBMC fraction enriched with plasma was collected and washed with PBS containing 0.1% human pooled serum and 1 mM UltraPure EDTA (0.5 M, pH 8, Life Technologies) at 190g for 15 min, RT, to separate the platelet rich plasma (PRP) from the PBMCs. The pellet containing PBMCs were then washed twice with cold PBS and resuspended in RPMI 1640 Dutch-modified culture medium supplemented with 1 mM pyruvate (Invitrogen), 2 mM glutamine (Invitrogen), 50 µg/mL gentamicin (Centrafarm).

The cell counts and PBMC composition were performed with the Sysmex-XN 450 hematology analyzer.

#### PRP collection and isolation of platelets

The PRP was collected from the plasma supernatant subsequently after Ficoll gradient separation. Collected PRP was diluted in PBS containing 0.1% human pooled serum and 1 mM UltraPure EDTA (0.5 M, pH 8, Life Technologies) and centrifuged at 190g for 15min. The supernatant containing PRP was collected and centrifuged at 2500g, 5minutes, 4°C and gently resuspended in previously described RPMI 1640 Dutch-modified culture medium. Platelet count was measured with the Sysmex – XN 450 hematology analyzer.

#### Monocyte isolation

Part of the PBMC fraction was used to isolate monocytes. CD14^+^ monocytes were isolated by negative selection by MACS pan-monocyte isolation kit (Miltenyi Biotec) according to manufacturer’s protocol. The purity and count were assessed by the Sysmex-XN 450 hematology analyzer.

#### Polymorphonuclear (PMN) cell isolation

After removing the PBMC fraction, neutrophil isolation was performed by hypotonic lysis. Briefly, leftover cells (PMNs and erythrocytes) were incubated with hypotonic lysis buffer (155 mM NH4Cl, 10 mM KHCO3) for 15 and 10 minutes on ice. Afterwards, PMNs were washed twice in PBS and resuspended in RPMI 1640 medium without phenol red (Gibco, 32404014) supplemented with 50μg/mL gentamicin (Centrafarm), 2mM glutamax (Gibco), and 1mM pyruvate (Gibco). Neutrophils were counted with Sysmex Hematoanalyzer and brought to 5×10^6^cells/ml concentration for the following experiments.

### Stimulation assays

#### PBMC stimulation

500.000 PBMCs per well were stimulated in duplicate with RPMI, LPS (10 ng/ml) (Sigma-Aldrich, E. coli serotype 055:B5, further purified as described^26^), Monosodium urate (MSU) crystals (300ug/ml), Pam3CYSK4 (Pam3Cys) 10 μg/mL (L2000, EMC Microcollections) for 24 hours in 96-wells round-bottom plates (Greiner) at 37 °C and 5% CO2. Supernatants were collected after 24 hours and stored at-80°C until measurements were performed.

7-day PBMC stimulation was performed with 500.000 cells per well with 10% human pooled serum with RPMI, LPS and PHA, without changing medium in 96-wells round-bottom plates (Greiner) at 37 °C and 5% CO2. Supernatants were collected after 7 days and stored at-80°C until measurements were performed.

#### Trained immunity assays

We used our previously published protocol of *in vitro* trained immunity assay^27^. Briefly, 100.000 monocytes per well were seeded to flat-bottom 96-wells plate for 1 hour at 37 °C and 5% CO2. After 1-hour, non-attached monocytes were removed during the collection of supernatants. Monocytes were trained with BCG SSI (750 ug/ml), β-glucan (2 ug/ml), and oxLDL 10 (ng/ml) in the presence of 10% human pooled serum (HPS) for 24h at 37 °C and 5% CO2. After 24 hours and 3 days media was refreshed to RPMI with 10% HPS. On day 6 the trained monocytes were restimulated with LPS (10 ng/ml) or Pam3Cys (10 ug/ml) for 24 hours. Supernatants were collected and stored at-80°C until measurements were performed.

#### Neutrophil stimulation

500.000 neutrophils per well were stimulated in duplicate in flat bottom 96-well plates (Corning, NY, USA) for 4 hours with culture medium only (as control), LPS (1 μg/ml), MSU (300 μg/ml), LPS and MSU (1 μg/ml and 300 μg/ml respectively), Pam3Cys (10 μg/ml), Nigericin (1 μM), Ethanol (dissolving agent for Nigericin) and Phorbol-12-myristate-13-acetate (PMA, Sigma) (50 nM) at 37°C with 5% CO2. After 4 hours, the plates were centrifuged for 8 minutes at 350g, and the supernatants were stored at-80°C until measurement.

### ROS assay

ROS production of neutrophils was determined by a luminol (5-amino-2,3, dihydro-1,4-phtalazinedione)-based luminescence assay. To opaque flat-bottom 96-well plates (Corning, NY, USA) 200.000 neutrophils per well were added and stimulated in quadruplicate with serum-opsonized zymosan, PMA, and culture medium as control. Chemiluminescence was measured at 142 second intervals for 1 hour at 37°C in a BioTek Synergy HTreader. The integral of relative luminescence units per second (RLU/sec) was measured.

### NETosis assays

#### NOX-dependent NET formation

200.000 neutrophils per well were seeded to flat-bottom 96-well plates (Corning, NY, USA) at 37°C for 20 minutes. After attachment of neutrophils, the supernatants were removed. The neutrophils were stimulated in quadruplicate with Nigericin (1 μM), Ethanol, PMA (50 μM) or culture medium as control for 3 hours at 37°C, 5% CO2.

#### NOX-independent NET formation

100.000.000 platelets were either kept unstimulated or activated with 156uM Thrombin Receptor Activator Peptide 6 (TRAP6, Sigma) for 30 minutes at 37°C, 5% CO2 in round bottom non-stick tubes (corning 352063, Fisher Scientific). In parallel, 200.000 neutrophils per well were attached to flat-bottom 96-well plates (Corning, NY, USA) at 37°C for 20 min. The neutrophils were stimulated in quadruplicate with unstimulated platelets, TRAP6-stimulated platelets or culture medium as control for 1 hour at 37°C, 5% CO2.

After both assays, the neutrophils were washed twice with warm PBS and NETs were treated by partial digestion in culture medium supplemented with 5 U/ml micrococcal nuclease (MNase, Worthington biochemical corporation) 20 min at 37°C, 5% CO2. MNase was inactivated by brief vortexing, and the neutrophils were pelleted by centrifugation. The supernatant containing partially digested NETs were kept at-80°C until measurement.

#### Measurement of DNA concentration in NETs with Sytox Orange

The DNA concentrations in the NET formation assay MNase-treated supernatants (NOX-dependent and NOX-independent formation assay) were quantified by adding 5 mM Sytox Orange Nucleic Acid Stain (Life Technologies) solution to undiluted sample. Fluorescence was measured with excitation and emission of 530/560nm using the BioTek Synergy HT multi-reader. All the measurements were done in duplicate.

### Circulating plasma protein and cytokine measurements

Cytokine and neutrophil granule-associated protein concentrations upon stimulation of PBMCs, monocytes and neutrophils, as well as circulating hsCRP were measured with commercially available Enzyme-linked Immunosorbent Assay (ELISA) kits according to instructions supplied by the manufacturer. For detailed information on the ELISA kits used please refer to **Supplementary Table 1**.

### Flow cytometry

Flow cytometric analyses were performed on EDTA whole blood samples. 2 panels were used to characterize monocyte and neutrophil phenotypes.

Red blood cell lysis was performed with BD Pharm Lyse buffer treatment for 15 minutes at room temperature (RT) in the dark. After washing with FACS buffer (1% BSA in PBS with 2mM EDTA) the pellet was resuspended in 100 µl of FACS Buffer and incubated with 10 µl Human TruStain FcX (Biolegend, San Diego, CA, USA) Fc block for 10 minutes. Cells were independently stained with the following two panels. For monocyte analysis, cells were stained using the following anti-human fluorochrome-conjugated antibodies: CD45, CD3, CD19, CD56, CD14, CD16, HLA-DR, CD11b, CD11c, CD41, CCR2, CCR5. For neutrophil analysis, the following anti-human fluorochrome-conjugated antibodies were used: CD45, a lineage cocktail containing CD3, CD56, CD19, CD20 and CD14, CD123, CD15, CD16, CD35, HLA-DR, CD62L, CD49d, CD10 and CD11b.

50 µl of cells were incubated with 50 ul of the either antibody mix for 30 min at RT in the dark. Then the cells were washed and resuspended in FACS buffer. Helix NiR viability dye was added to the neutrophil panel for 15 min at RT in the dark. Cell populations and expression of markers were measured using a CytoFlex cytometer (Beckman Coulter, Brea, USA) which underwent daily quality control to correct for variation in laser settings. Flow Minus One controls were assessed for all markers for which MFI was assessed to set the correct gates. For a full overview of used antibodies see **Supplementary Table 2**.

Manual gating of the monocyte panel was performed by selecting for single cells and CD45+ immune cells, B cells, T cells and NK cells were removed based on the expression of CD19, CD3 and CD56. CD19-CD3-CD56-cells were used for monocyte gating; monocytes were identified based on the expression of CD14 and CD16 and HLA-DR. Monocyte subsets (classical, intermediate and nonclassical) were determined based on CD14 and CD16 expression and the exact gates were put based on HLA-DR and CCR5 expression (highest on intermediate monocytes), and CCR2 expression (highest on nonclassical monocytes) (**Supplementary Figure 1**).

Manual gating of the neutrophil panel was performed by removing doublets and debris and selecting for live granulocytes among CD45+ cells. Then, neutrophils were identified based on CD15 and CD16 surface expression. After excluding eosinophils and basophils, we characterized the maturation and activation level of neutrophils by CD49d, CD10, CD66b, HLA-DR, CD16 and CD62L (**Supplementary Figure 2**)

### Statistical analysis

Distribution of data was assessed with the Shapiro-Wilk test. If the data did not follow a normal distribution, it is shown as median and interquartile range. Mann-Whitney U test was used to compare DNMT3A and No CHDM groups. P<0.05 is considered statistically significant and is indicated with an asterisk in tables. Statistical analyses were performed by using Graphpad version 9 and SPSS.

## Results

### Baseline characteristics did not differ among groups

Initially we included 36 individuals, 15 of whom had a DNMT3A CHDM, 15 without CHDMs and 6 with TET2 CHDM based on 2016 sequencing. However, based on the 2023 sequencing, one of the individuals without CHDMs gained a DNMT3A mutation, and for one individual there were not enough reads to confirm the absence of mutations. Two of the TET2 CHDMs (with very small VAFs, i.e. ∼0.01) could not be repeated, therefore those individuals were moved to No CHDM, and one individual with TET2 mutation developed a DNMT3A mutation. Thus, we excluded the remaining 3 individuals with TET2 mutations. And the final groups in the current study included 15 individuals who did not have any known CHDMs and 17 had DNMT3A mutations. For the visual representation of inclusion please refer to **Figure 1**. The baseline characteristics of the participants are listed in **Table 1**. The median age was 75 years and BMI of 30 kg/m^2^. All participants had Western European ancestry and 47% were male. There were no statistical differences in the baseline characteristics of the two groups.

### Majority of the DNMT3A mutations persisted in time

We identified DNMT3A CHDMs in 17 individuals. 2 of these mutations were newly identified in individuals that previously did not have CHDMs. Of the previously identified clone trajectories 8 clones grew, 2 remained stable and 7 shrank in size, based on the VAFs measured in 2016 and 2023. Clone characteristics and dynamics of the mutations are shown in **Table 2**.

**Table 2:** Clone characteristics.

| Position | Coding DNA annotation | Protein amino acid annotation | Amino acid change | VAF 2016 | VAF 2023 |
| --- | --- | --- | --- | --- | --- |
| 25457243 | c.2644C>T | p.Arg882Cys | R -> C (882) | 18.2 | 25.7 |
| 25463182 | c.2311C>T | p.Arg771Ter | R -> [STOP] (771) | 11.4 | 6.9 |
| 25457161 | c.2726T>G | p.Phe909Cys | F -> C (909) | 4.8 | 6.1 |
| 25462020 | c.2387G>T | p.Gly796Val | G -> V (796) | 22.9 | 22.3 |
| 25464537 | c.1976G>A | p.Arg659His | R -> H (659) | 3.3 | 5.0 |
| 25463521 | c.2161A>T | p.Lys721Ter | K -> [STOP] (721) | 34.5 | 45.0 |
| 25457242 | c.2645G>A | p.Arg882His | R -> H (882) | 7.8 | 5.9 |
| 25467083 | c.1792C>T | p.Arg598Ter | R -> [STOP] (598) | 3.5 | 5.5 |
| 25463289 | c.2204A>G | p.Tyr735Cys | Y -> C (735) | 6.5 | 4.7 |
| 25457242 | c.2645G>A | p.Arg882His | R -> H (882) | 11.3 | 10.6 |
| 25457246 | c.2641delA | p.Ser881Alafs*25 | [STOP] AA 905 | 5.1 | 7.8 |
| 25470498 | c.976C>T | p.Arg326Cys | R -> C (326) | 16.7 | 14.0 |
| 25463181 | c.2312G>A | p.Arg771Gln | R -> Q (771) | 2.8 | 3.5 |
| 25463554 | c.2128T>A | p.Cys710Ser | C -> S (710) | 6.0 | 3.6 |
| 25468919 | c.1444G>T | p.Glu482Ter | E -> [STOP] (482) | 3.3 | 3.3 |
| 25463568 | c.2114T>C | p.Ile705Thr | I -> T (705) | 6.1 | 4.4 |
| 25457242 | c.2645G>A | p.Arg882His | R -> H (882) | 0 | 0.6 |

### DNMT3A mutations did not affect the leukocyte composition

In order to study if the presence of DNMT3A mutations alter the circulating leukocyte composition we measured leukocyte number and differentiation. Absolute counts and percentages of circulating leukocytes did not differ among individuals with DNMT3A mutations or without CHDMs (**Figure 2A-B**). Also, there was no difference in hsCRP, an indicator of systemic inflammation (data not shown).

**Figure 2:**
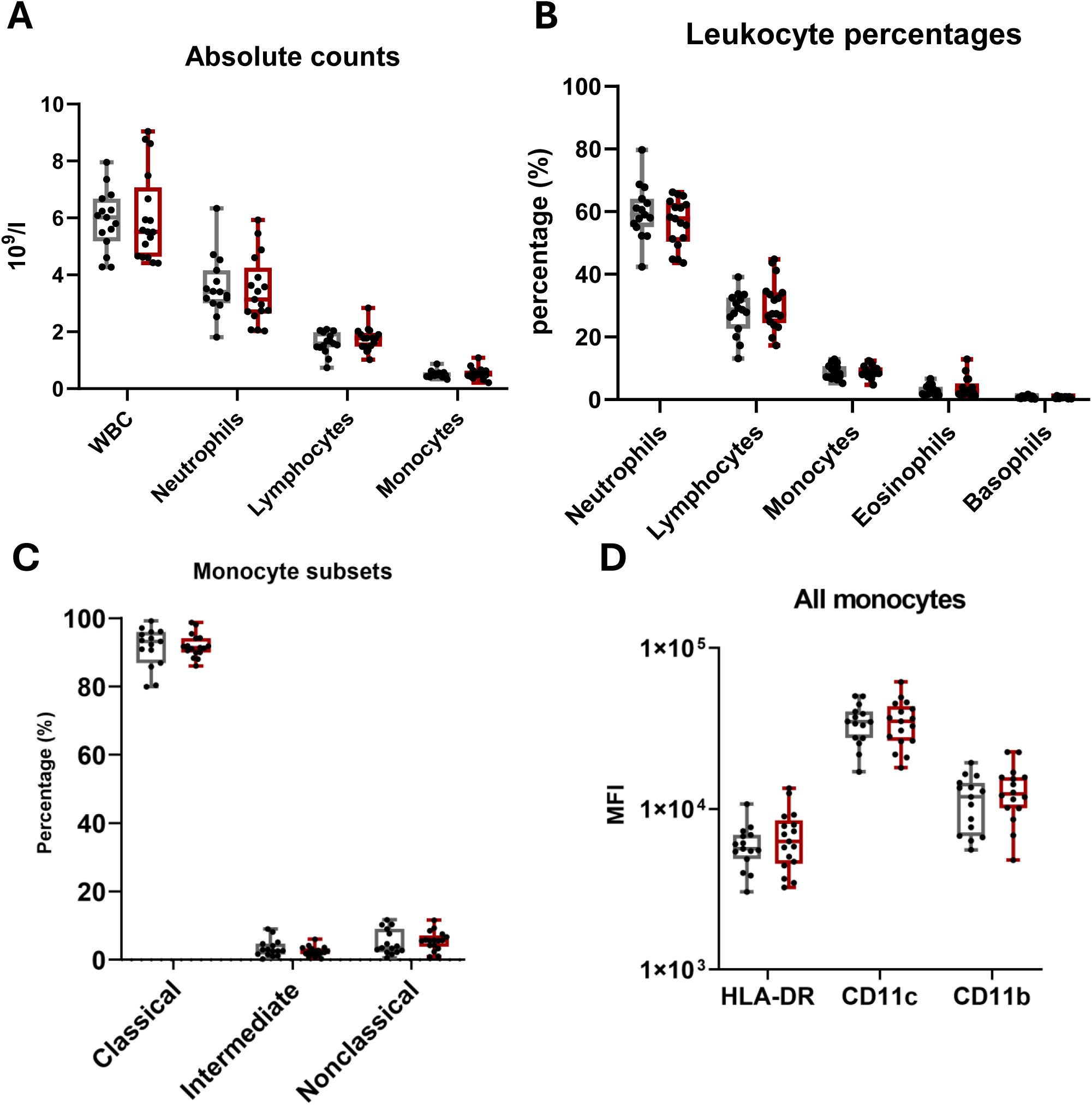
Leukocyte counts and monocyte phenotype, No CHDM: Gray, DNMT3A: Red. Absolute counts and percentages of monocytes measured with Sysmex Hematoanalyzer. Percentages of monocyte subsets and some key activation markers measured with multicolor flow cytometry. WBC: White Blood Cell, HLA: Human Leukocyte Antigen DR isotype, CD: Cluster of Differentiation

### Monocyte subsets or activation status did not differ in individuals with DNMT3A mutations

To investigate whether DNMT3A mutations were associated with changes in the monocyte phenotype and subsets we performed multicolor flow cytometry. We did not observe any difference in the percentages of classical, intermediate, or non-classical monocytes among DNMT3A carriers or individuals without CHDMs (**Figure 2C**).

In total monocytes as well as subsets, there were no difference in activation markers HLA-DR, CCR2, CCR5, CD11b, CD11c and CD41, both with regard to the percentage of cells with these markers, as well as for MFI, between individuals without CHDMs and with DNMT3A mutations (**Figure 2D**).

### The effect of DNMT3A mutations on PBMC ex vivo cytokine production capacity

To assess whether monocytes are functionally distinct in individuals with DNMT3A mutations or without CHDMs we stimulated isolated PBMCs *ex vivo* for 24 hours with various PRR ligands, as previously done for the total cohort of 297 individuals^6^. For the majority of the cytokines, there was a trend for lower cytokine production in individuals with DNMT3A mutations, in line with our previous observations in the entire 300 OB cohort, although this did not reach statistical significance, apart from IL1RA production upon MSU stimulation, that was significantly lower in individuals with DNMT3A mutations (**Figure 3D**).

**Figure 3:**
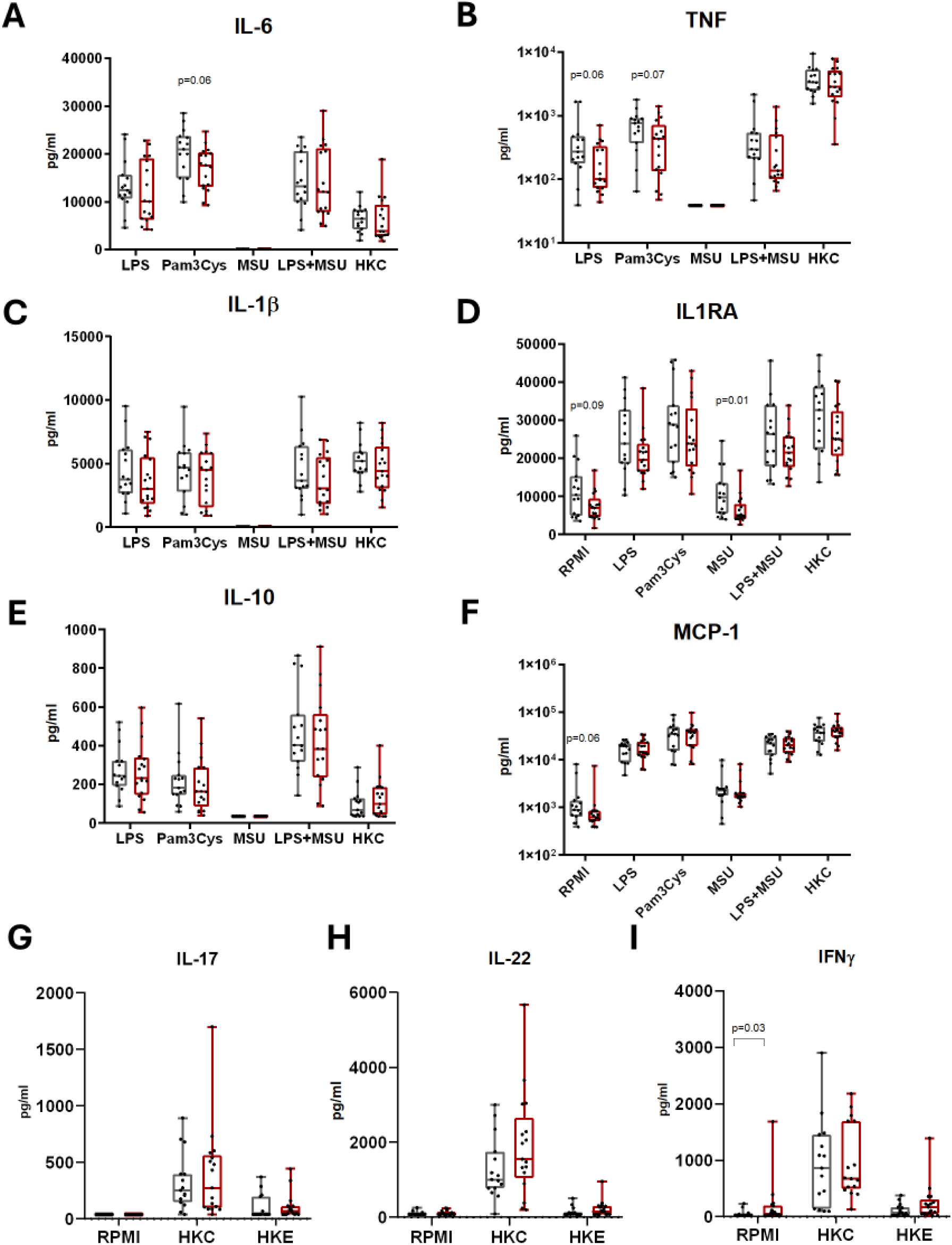
Ex vivo cytokine production capacity of PBMCs, No CHDM: Gray, DNMT3A: Red. Panels A-F demonstrate cytokine production upon 24-hour stimulation of PBMCs with the following stimuli LPS(Lipopolysaccharide), Pam3Cys (Pam3CYSK4), MSU (Mono sodium urate), HKC (Heat-killed Candida albicans). Panels G-I demonstrate cytokine production upon 7-day stimulation of PBMCs with HKC (Heat-killed Candida albicans) and HKE (Heat-killed Escherichia coli). IL: Interleukin, TNF: Tumor necrosis factor, MCP-1: Monocyte Chemoattractant Protein-1, IFNγ: Interferon gamma

### The effect of DNMT3A mutations on trained immunity

We subsequently set out to test our hypothesis that trained immunity induction is augmented in monocytes from individuals with DNMT3A CHDMs. We induced trained immunity by exposing isolated monocytes for 24 hours to three different triggers of trained immunity, β-glucan, BCG and oxLDL. After 6 days of differentiation into macrophages we restimulated the cells with LPS and Pam3Cys and measured cytokine production.

The trained immunity capacity is calculated as a fold change cytokine production over the background control (RPMI). For detailed investigation of the monocyte cytokine production and trained immunity capacity we show both raw cytokine production values as well as the fold change. Similar to the PBMC *ex vivo* cytokine production capacity, there was a general trend for lower cytokine production in the macrophages in individuals with DNMT3A mutations (**Figure 4A, C, E**). Particularly when monocytes were untrained (i.e. only exposed to RPMI) and restimulated with TLR ligands there was a trend for lower cytokine production (**Figure 4A** TNF production of LPS restimulated RPMI, p=0.07; **Figure 4E** IL1RA production of Pam3Cys restimulated RPMI, p=0.04).

**Figure 4:**
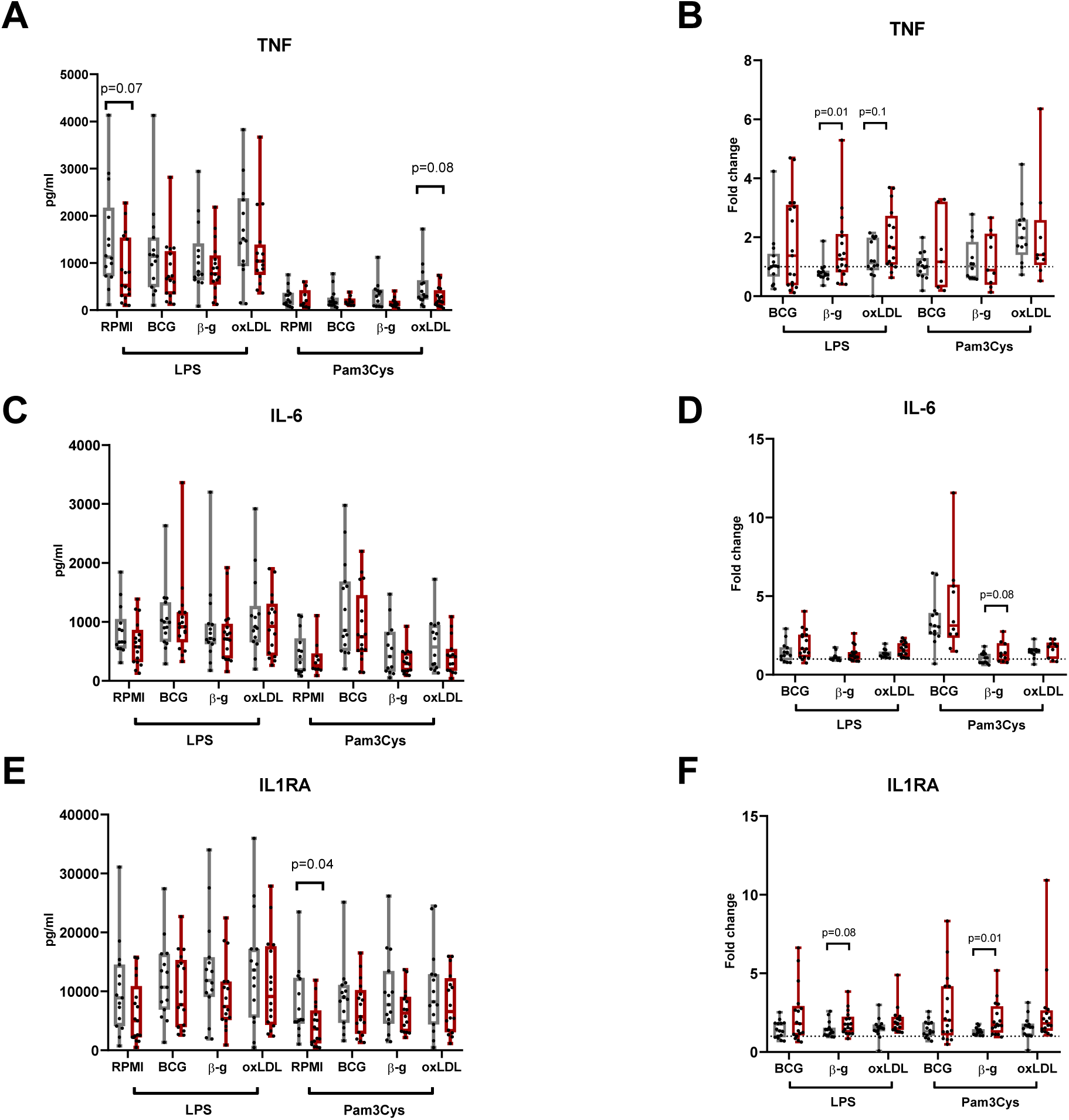
Monocyte training. No CHDM: Gray, DNMT3A: Red. Graphs on the left column(A, C, E) show raw values of cytokine production capacity with training stimuli BCG (Bacillus Calmette– Guérin SSI), β-g (β-glucan), oxLDL (oxidized low-density lipoprotein), and RPMI as background control restimulated with either LPS or Pam3Cys. Graphs on the right column (B, D, F) show fold change cytokine production over RPMI. Dashed line at fold change 1.

In contrast, the capacity to build trained immunity, calculated as fold change in cytokine production after 24 hour exposure to the training stimuli, compared to the un-trained control situation, was higher in individuals with DNMT3A mutations for several cytokines and training stimuli (**Figure 4**). Specifically, β-glucan (p=0.01) and oxLDL (p=0.1) training induced higher TNF fold change after LPS restimulation (**Figure 4B**); β-glucan training resulted in a higher IL-6 fold change after Pam3Cys restimulation (p=0.08) (**Figure 4D**); and β-glucan training induced higher IL1RA fold change with both LPS (p=0.08) and Pam3Cys (p=0.01) restimulation (**Figure 4F**).

### DNMT3A driven CH did not associate with changes in the adaptive immune response

It has been shown that clonal hematopoiesis predominantly skews the HSPCs towards myelopoiesis. Thus, the effects of CHDMs on lymphocyte function are less well characterized. To provide an in-depth immune characterization, we measured some key adaptive cytokines; IL-17, IL-22 and IFNy upon stimulation with heat-killed *C. albicans* and *E. coli* for 7 days (**Figure 3G-I**). Except for higher IFNy production on the baseline for individuals with DNMT3A mutations, we did not observe any statistically significant differences among the groups (**Figure 3I**).

### The effects of DNMT3A mutations on neutrophil phenotype and function

We phenotypically characterized neutrophils from whole blood with a multicolor flow cytometry approach. We assessed a panel of maturation and activation markers. Neutrophils from individuals with DNMT3A mutations were characterized by a higher expression of the CD10 maturation marker, without changes in the percentage of cells expressing CD10 (**Table 3**). We observed that individuals with DNMT3A mutations had higher percentage of neutrophils expressing HLA-DR, although this did not reach statistical significance (p=0.1). There were no differences in expression of the other surface markers.

**Table 3:** Neutrophil flow cytometric characterization.

| FCM Mature PMN | No CHDM (n=15) |  | DNMT3A (n=17) |  | P-Value |
| --- | --- | --- | --- | --- | --- |
|  | Mean | SD | Mean | SD |  |
| % of neutrophils | 99.7 | 0.2 | 99.7 | 0.2 | 0.8 |
| CD10 MFI | 63287 | 19245 | 77250 | 21588 | **0.04** |
| CD10 % | 98.0 | 1.5 | 98.7 | 1.0 | 0.2 |
| CD66b MFI | 15127 | 3628 | 16815 | 4805 | 0.4 |
| CD15 MFI | 36551 | 9582 | 43853 | 17703 | 0.3 |
| CD11b % | 1.0 | 0.0 | 1.0 | 0.0 | >0.9 |
| CD11b MFI | 14333 | 5005 | 19002 | 7518 | 0.1 |
| CD35 % | 100 | 1 | 100 | 1 | 0.6 |
| CD35 MFI | 12735 | 4681 | 14021 | 5956 | 0.7 |
| CD62 % | 99.7 | 0.2 | 99.3 | 0.9 | **0.01** |
| CD62L MFI | 61391 | 25071 | 62173 | 21816 | 0.9 |
| HLA-DR % | 32.5 | 37.1 | 60.3 | 39.4 | 0.1 |
| HLA-DR % | 3699 | 3228 | 3730 | 3118 | 0.3 |

We assessed the degranulation capacity of neutrophils based on primary (MPO, NE) and secondary granule (s100A8/9, NGAL) contents with ELISA. Neutrophils from individuals with DNMT3A mutations had significantly lower MPO release upon TLR2 signaling via Pam3Cys stimulation. We did not identify differences in the concentrations of any other granule marker (**Table 4**).

**Table 4:** Neutrophil ex vivo degranulation capacity.

|  | PMN 4h | No CHDM (n=15) |  | DNMT3A (n=17) |  | p-value |
| --- | --- | --- | --- | --- | --- | --- |
|  |  | Mean | SD | Mean | SD |  |
| **NGAL**<br>**(ng/ml)** | RPMI | 11.0 | 4.6 | 10.1 | 4.6 | 0.65 |
|  | LPS | 26.2 | 11.7 | 24.2 | 14.4 | 0.65 |
|  | Pam3Cys | 62.9 | 23.0 | 58.1 | 17.1 | 0.65 |
|  | MSU | 23.8 | 10.6 | 23.1 | 20.6 | 0.14 |
|  | LPS+MSU | 29.5 | 10.2 | 32.0 | 24.8 | 0.18 |
|  | Nigericin | 48.5 | 17.6 | 44.0 | 23.8 | 0.22 |
|  | Ethanol | 20.3 | 13.8 | 14.0 | 8.6 | **0.05** |
|  | PMA | 198.5 | 44.5 | 206.6 | 37.5 | 0.97 |
| **MPO**<br>**(ng/ml)** | RPMI | 142.9 | 91.2 | 130.1 | 91.8 | 0.68 |
|  | LPS | 208.7 | 146.8 | 159.5 | 90.7 | 0.41 |
|  | Pam3Cys | 144.9 | 80.5 | 85.3 | 37.1 | **0.02** |
|  | MSU | 241.2 | 117.2 | 225.8 | 159.4 | 0.39 |
|  | LPS+MSU | 291.9 | 128.8 | 277.3 | 177.1 | 0.6 |
|  | Nigericin | 455.7 | 151.8 | 398.4 | 132.0 | 0.28 |
|  | Ethanol | 190.5 | 134.9 | 134.9 | 113.8 | 0.18 |
|  | PMA | 596.8 | 292.2 | 612.5 | 347.5 | 0.82 |
| **S100A8/9**<br>**(ng/ml)** | RPMI | 291.8 | 117.3 | 254.1 | 99.1 | 0.37 |
|  | LPS+MSU | 1395.7 | 604.5 | 1100.9 | 502.9 | 0.2 |
|  | Pam3Cys | 1931.6 | 866.9 | 1530.5 | 350.9 | 0.16 |
|  | MSU | 1846.2 | 679.4 | 1927.3 | 1422.8 | 0.37 |
|  | LPS+MSU | 1839.0 | 563.9 | 1776.3 | 1159.0 | 0.26 |
|  | Nigericin | 48.5 | 17.6 | 44.0 | 23.8 | 0.22 |
|  | Ethanol | 1528.9 | 674.5 | 1124.6 | 511.2 | **0.04** |
|  | PMA | 8080.0 | 2278.6 | 7441.5 | 2026.9 | 0.55 |
| **IL-8 (pg/ml)** | RPMI | 23.5 | 0 | 23.5 | 0 | >0.99 |
|  | LPS | 31.3 | 17.9 | 24.3 | 3.4 | 0.09 |
|  | PMA | 191.6 | 81.7 | 241.8 | 156.8 | 0.5 |

Neutrophils can expel their DNA in the form of extracellular traps, which contributes to atherosclerosis development and plaque destabilization. In addition to the DNA content, these neutrophil extracellular traps (NETs) contain nuclear, cytoplasmic and granular proteins^22^. We quantified the amount of DNA released during the NOX-dependent and –independent NET formation assays with a Sytox based assay. The presence of DNMT3A mutations was not associated with changes in the concentration of DNA released during NET formation (data not shown).

An important function of neutrophils is production of reactive oxygen species (ROS), involved in antimicrobial host defense and inflammation. We measured ROS production capacity of neutrophils upon stimulation with opsonized zymosan and PMA. There were no statistically significant changes in the ROS production capacity of neutrophils from individuals without CHDMs or DNMT3A mutations (data not shown).

## Discussion

In the present study, we aimed to explore two potential immunological mechanisms that could contribute to the increased cardiovascular risk in patients with clonal hematopoiesis due to CHDMs in the *DNMT3A* gene. First, we showed that the presence of a DNMT3A CHDM is associated with an increased trained immunity response. Secondly, we explored in detail how DNMT3A CHDMs affected neutrophil phenotype and function, and showed that individuals with DNMT3A mutations had more CD10^+^ mature neutrophils and lower MPO release after TLR2 stimulation. These findings offer exciting new immunological pathways that can contribute to the link between CHIP and CVD.

We focused specifically on DNMT3A CHDMs, since mutations in the DNMT3A gene are the leading drivers of clonal hematopoiesis, they are associated with ASCVD^11^, and because of sequencing covered the entire DNMT3A gene, in contrast to the other genes involved in CH. We selected individuals from our 300 OB study, based on the sequencing results from 2016, and repeated this sequencing in 2023^6^. Clone size of TET2-driven CH is shown to grow exponentially with age, whereas DNMT3A clone growth dynamics are slower and mostly stable^28^. Approximately 60% of the clone trajectories identified in our cohort either grew or remained static.

One of the two main aims of this study was to explore the hypothesis that CH is associated with an increased tendency to develop trained immunity. Trained immunity refers to the immunological phenomenon that innate immune cells, such as monocytes, can build a long-term hyperinflammatory phenotype after brief stimulation, e.g. with micro-organisms, but also with endogenous atherogenic molecules, such as oxLDL^29^, high glucose concentrations^16^, or catecholamines^30^. This is established through rewiring of key metabolic pathways, and through epigenetic reprogramming^13^. Accumulating experimental evidence irrefutably showed that trained immunity can accelerate the development of atherosclerosis^16,18^. We argued that *DNMT3A*-related CH could modulate the susceptibility to mount trained immunity responses for two reasons. First, *DNMT3A* encodes for a histone methyltransferase and trained immunity is regulated by epigenetic processes, including histone methylation, and DNA methylation^31,32^. Secondly, it is known that IL-1β signaling is involved in the development of the trained phenotype, at least for trained immunity induced by oxLDL and by β-glucan^14,15^. Because there are indications that DNMT3A deficiency increases IL-1β expression^11,12^, this could be another mechanism linking CH and trained immunity.

We first recapitulated our previous findings from the 300 OB cohort of a lower *ex vivo* cytokine production capacity of PBMCs from individuals with DNMT3A mutations^6^. Interestingly, a recent single cell RNA sequencing study in unstimulated monocytes from individuals with heart failure revealed a higher proinflammatory cytokine expression at baseline^12^.

In line with our hypothesis, monocytes from individuals with DNMT3A mutations were more amenable to be trained by β-glucan, which reached significance for TNF production upon TLR4 restimulation and IL1RA production upon TLR2 restimulation. Also, there was a trend for higher TNF production in LPS restimulated oxLDL trained monocytes however this did not reach statistical significance (p=0.1). This could partially be explained by the fact that oxLDL is not a stimulus as potent as β-glucan. In conclusion, baseline cytokine production capacity of monocytes from individuals with DNMT3A was lower, whereas capacity to build trained immunity was higher compared to individuals without CHDMs. This finding nicely fits the framework predicting trained immunity responses that was recently described in a cohort of 323 healthy individuals that were trained with BCG vaccination *in vivo*^33^. The authors showed that 213 individuals had a trained immunity response, whereas 78 individuals were categorized as non-responders. Interestingly, individuals with the strongest BCG-induced trained immunity responses, were characterized by low cytokine production at baseline (i.e. before BCG administration) and low chromatin accessibility at genes involved in innate immunity^33^. The lower baseline cytokine production capacity in our study in individuals with DNMT3A CHDMs could potentially be due to intrinsic effects of the DNMT3A mutation on cytokine production, or could be by the fact that individuals with CHDMs already have elevated levels of systemic inflammation as we previously shown^6^, that itself induces a state of immune-tolerance. Future studies are needed to investigate the epigenetic landscape of the DNMT3A mutated cells to understand how this affects trained immunity. In addition, single cell sequencing studies should be performed to investigate the individual cellular effects of DNMT3A CHDMs on cytokine production and on trained immunity.

The second main aim of our study was to investigate in detail how DNMT3A CHDMs affect the phenotype and function and neutrophils. There is accumulating evidence that neutrophils are important regulators of atherosclerosis development^21,22^. In addition, we recently showed that the atherosclerosis accelerating effect of high fat diet-induced trained immunity is dependent on neutrophil activation. Up to now, information about how CHDMs affect neutrophil function is scarce, and mainly stems from CHDMs in TET2 and JAK^19,20^. We did not find relevant effects on neutrophil surface marker expression or protein, NET, and ROS release, except a higher expression of CD10. CD10^-^ immature neutrophils have been found to associate with increased inflammatory markers and cardiac damage in patients with acute myocardial infarction^34^.

In a TET2 mutant mouse model, neutrophils were shown to have increased primary granule content marked by MPO and NE quantity^19^. In contrast, our findings illustrate that MPO quantity upon Pam3Cys stimulation was lower in the neutrophils from individuals with DNMT3A mutations. This could hint to a gene specific effect in clonal hematopoiesis and should be investigated in future studies. The same study demonstrated that the NET architecture from TET2 mutant neutrophils was altered. We measured the amount of DNA released during NOX-dependent and –independent NET formation. The released DNA concentrations were comparable among the two groups, alas we were not able to characterize the NET architecture.

A limitation of our study is the small sample size, which prevented us from reaching robust conclusions for some immunological parameters. However, we selected these individuals from a larger cohort in which we previously also demonstrated a lower cytokine production capacity in individuals with CHDM, showing robust internal validity. A second limitation is that our cohort consisted of individuals with Western European ancestry, thus our findings cannot be extended to diverse ethnic groups.

A strength of our study is the in-depth immune characterization including phenotypic and functional characterization of monocytes and neutrophils, and adaptive immune responses. In addition to the extensive immunological characterization, we strived for a balanced distribution of sex, age and BMI between groups to minimize potential confounding factors. To the best of our knowledge, this is the first study that investigated the association between trained immunity and clonal hematopoiesis in individuals with obesity.

In conclusion, we show that in individuals with obesity, DNMT3A mutations are associated with *lower ex vivo* cytokine production capacity and a heightened susceptibility to develop a trained immunity phenotype in response to stimuli. Future studies are necessary to unravel the underlying mechanism of this association and the consequences for the development of atherosclerotic CVD.

## Acknowledgements

The authors thank H. Lemmers, and H. Dijkstra for their help with ELISAs.

## Data availability

The anonymized data underlying this study is available upon reasonable request to the corresponding author.

## Supplementary Information

**Supplementary Table 1:** ELISA kits used in this study.

| Product | Product number | Manufacturer |
| --- | --- | --- |
| Human IL-1β DuoSet ELISA | DY201 | Bio-Techne/R&D |
| Human IL-1RA DuoSet ELISA | DY280 | Bio-Techne/R&D |
| Human IL-6 DuoSet ELISA | DY206 | Bio-Techne/R&D |
| Human IL-10 DuoSet ELISA | DY217B | Bio-Techne/R&D |
| Human TNF DuoSet ELISA | DY210 | Bio-Techne/R&D |
| Human hsCRP ELISA | DY1707 | Bio-Techne/R&D |
| Human IL-8 DuoSet ELISA | DY208 | Bio-Techne/R&D |
| Human Neutrophil Elastase/ELA2 DuoSet ELISA | DY9167-05 | Bio-Techne/R&D |
| Human S100A8/S100A9 Heterodimer DuoSet ELISA | DY8226-05 | Bio-Techne/R&D |
| Human Myeloperoxidase DuoSet ELISA | DY3174 | Bio-Techne/R&D |
| Human Lipocalin-2/NGAL DuoSet ELISA | DY1757 | Bio-Techne/R&D |

**Supplementary Table 2:** Antibodies used for flow cytometry.

| Antibodies | Fluorochrome | Clone | Company | Identifier |
| --- | --- | --- | --- | --- |
| Anti-human CD16 | FITC | 3G8 | Biologend | Cat# 302006 RRID AB_314206 |
| Anti-human HLA-DR | PE | immu-357 | Beckman Coulter | Cat# IM1639U RRID AB_2876782 |
| Anti-human CD62L | PEdazzle584 | DREG-56 | Biologend | Cat# 304842 RRID AB_2565874 |
| Anti-human CD49d | PECy5.5 | 9F10 | Biologend | Cat# 304312 RRID AB_10641699 |
| Anti-human CD10 | PC7 | HI10a | Biologend | Cat# 312213 RRID AB_2146549 |
| Anti-human lineage cocktail (CD3, CD14, CD19, CD20, CD56) | APC | UCHT1 ; HCD14 ; HIB19; 2H7; HCD56 | Biologend | Cat# 348703 RRID: N/A |
| Anti-human CD66b | APC-700 | G10F5 | Biologend | Cat# 305114 RRID AB_2566038 |
| Anti-human CD15 | APC-Cy7 | MEM-166 | Biologend | Cat# 323047 RRID AB_2750189 |
| Anti-human CD123 | BV421 | 6H6 | Biologend | Cat# 306018 RRID AB_10962571 |
| Anti-human CD45 | BV510 | HI30 | Biologend | Cat# 304036 RRID AB_2561940 |
| Anti-human CD35 | BV650 | E11 | BD Biosciences | Cat# 744277 RRID AB_2742115 |
| Anti-human CD11b | BV785 | ICRF44 | Biolegend | Cat# 301346 RRID AB_2563794 |
| Anti-human CD11c | PEDazzle584 | BU15 | Biolegend | Cat# 337227 RRID AB_2564548 |
| Anti-human CD3 | PC5.5 | UCHT1 | Biolegend | Cat#300410 RRID AB_314064 |
| Anti-human CD14 | PC7 | 61D3 | eBioscience | Cat#25-0149 RRID AB_1582276, |
| Anti-human CD56 | APC | N901 | Beckman Coulter | Cat# IM2474, RRID AB_130791 |
| Anti-human CD19 | AF700 | HIB19 | Biolegend | Cat: 302226, RRID AB_493751 |
| Anti-human CD41 | APC-Cy7 | HIP8 | Biolegend | Cat: 303716, RRID AB_10897646 |
| Anti-human CCR2 | BV421 | 48607 | BD Biosciences | Cat: 564067, RRID AB_2738573 |
| Anti-human CCR5 | BV650 | 3A9 | BD Biosciences | Cat: 564999, RRID: AB_2739037 |
| Brilliant stain buffer | - | - | BD Biosciences | Cat: 563794, RRID: N/A |
| Helix NP™ NIR | - | - | Biolegend | Cat# 425301, RRID, N/A |

**Supplementary Figure 1:**
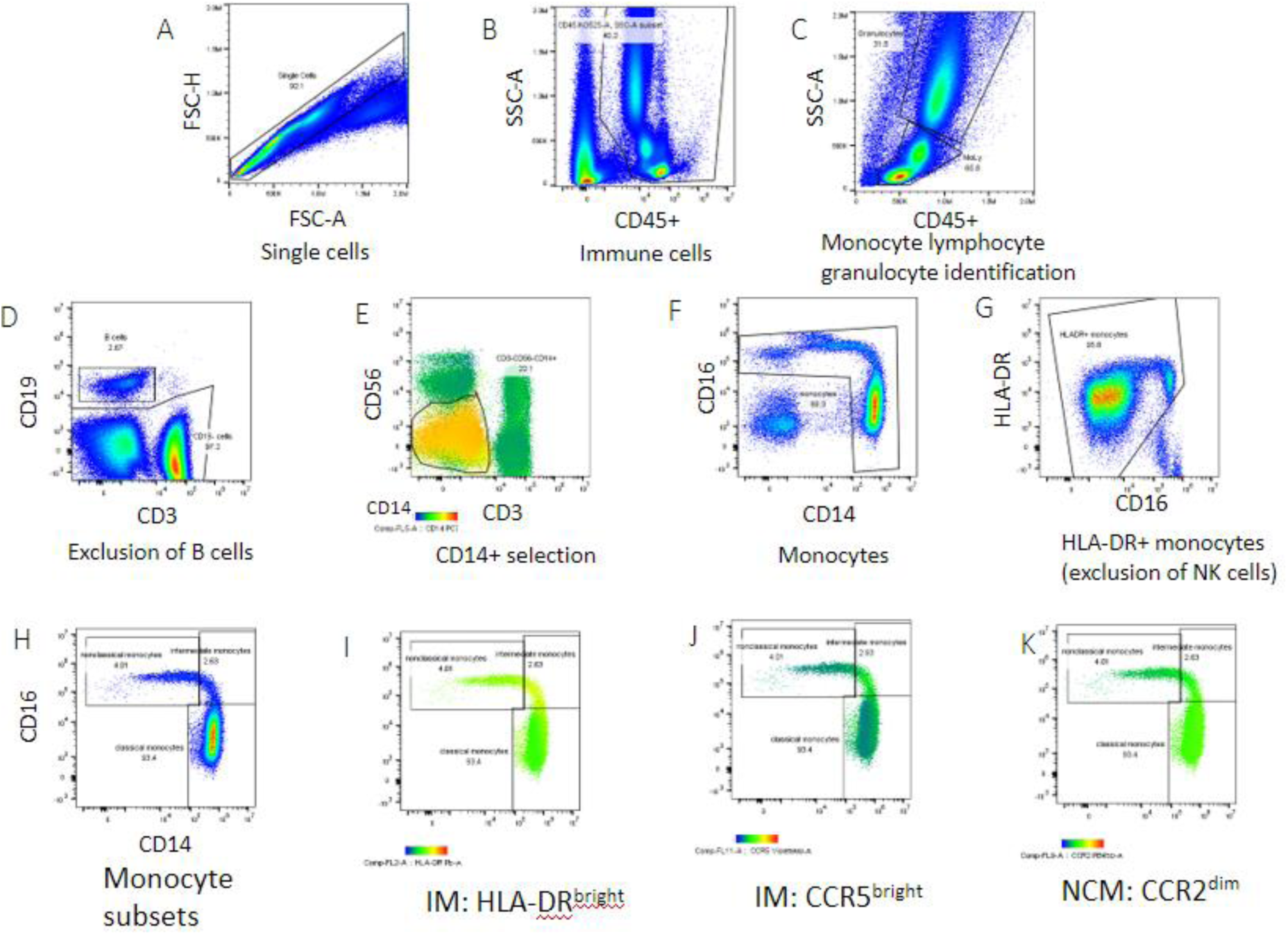
Gating strategy defining monocyte population using flow cytometry. Manual gating strategy for monocyte subsets: single cells (A) and CD45+ immune cells(B). Gating for monocytes, lymphocytes and granulocytes based on forward and side scatter(C). We excluded B-cells based on high expression of CD19(D). Thereafter, monocytes were identified based on the expression of CD14 and CD16 (E-F) HLA-DR+ monocytes were selected to eliminate NK cells (G). Monocyte subsets (classical, intermediate and nonclassical) were determined based on CD14 and CD16 expression and the exact gates were put based on HLA-DR and CCR5 expression (highest on intermediate monocytes), and CCR2 expression (highest on nonclassical monocytes) (H-K).

**Supplementary Figure 2:**
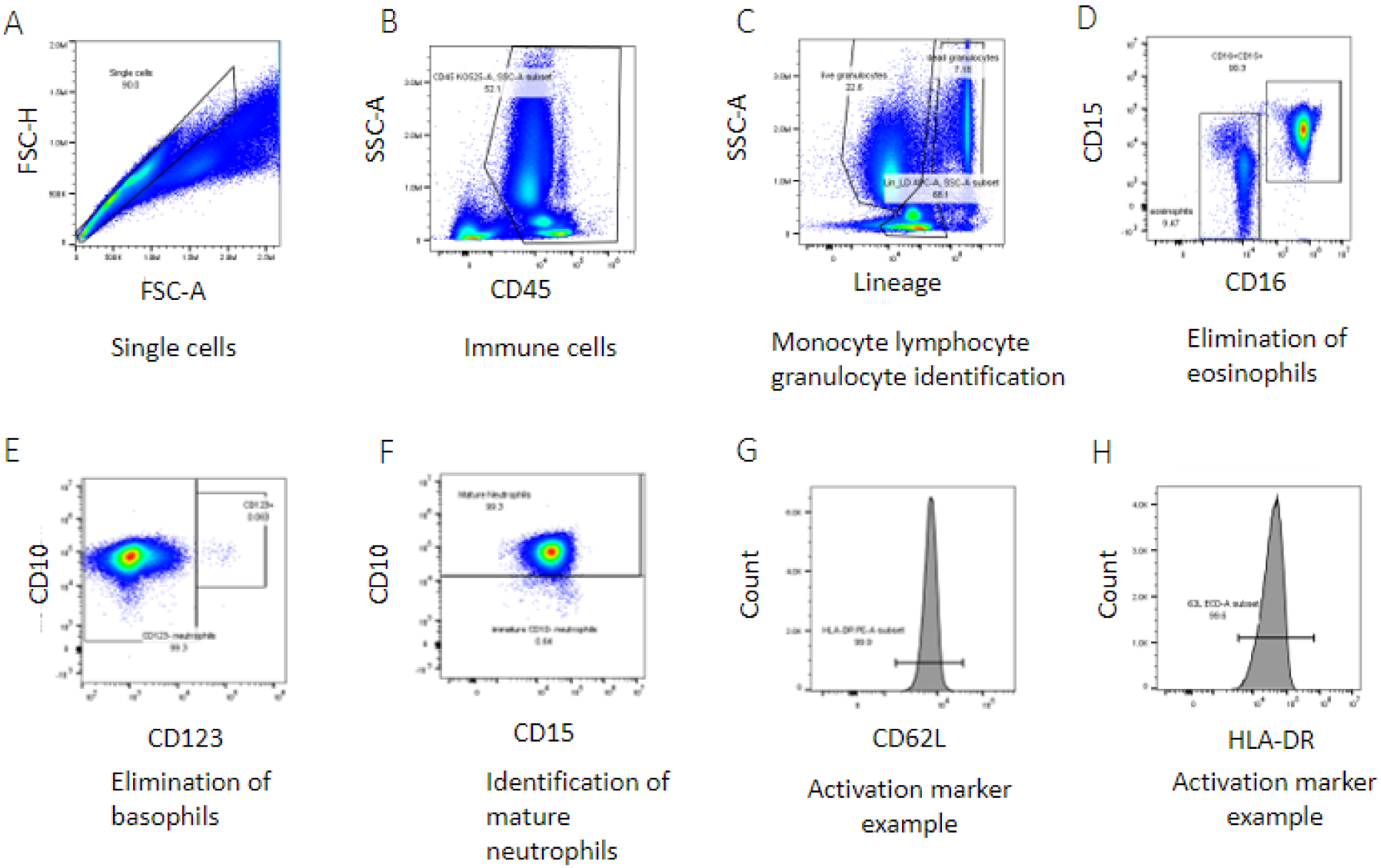
Gating strategy for defining neutrophil population using flow cytometry. Manual gating strategy for neutrophil sub-analysis: gating for (A) single cells and (B) CD45+. Then granulocytes were selected based FSC/SSC (C). Eosinophils and basophils were eliminated based on low CD16 expression, and high CD123 expression respectively (D-E). Mature neutrophils were determined based on CD10 expression. (F) Neutrophils were further analyzed for their median fluorescent intensity of activation markers HLA-DR (G) and CD62L (H

